# Lopinavir-Ritonavir in treatment of COVID-19: A dynamic systematic benefit-risk assessment

**DOI:** 10.1101/2020.05.27.20114470

**Authors:** Vicki Osborne, Miranda Davies, Samantha Lane, Alison Evans, Jacqueline Denyer, Sandeep Dhanda, Debabrata Roy, Saad Shakir

## Abstract

**Background:** COVID-19 is an ongoing, global public health crisis for which safe and effective treatments need to be identified. The benefit-risk balance for use of lopinavir-ritonavir in COVID-19 needs to be monitored on an ongoing basis, therefore a systematic benefit-risk assessment was designed and conducted. A key objective of this study was to provide a platform for a dynamic systematic benefit-risk evaluation; although initially this evaluation is likely to contain limited information, it is required due to the urgent unmet public need. Importantly it allows additional data to be incorporated as it becomes available, and re-evaluation of the benefit-risk profile.

**Methods:** A systematic benefit-risk assessment was conducted using the Benefit-Risk Action team (BRAT) framework. The exposure of interest was lopinavir-ritonavir treatment in COVID-19 compared to standard of care, placebo or other treatments. A literature search was conducted in PubMed and EmBase to identify peer-reviewed papers reporting clinical outcomes. Two clinicians constructed a value tree and ranked key benefits and risks in order of considered clinical importance.

**Results:** In comparison to standard of care, data for several key benefits and risks were identified for lopinavir-ritonavir. Time to clinical improvement was not significantly different for lopinavir-ritonavir in comparison to standard of care (HR=1.31, 95% CI:0.95, 1.80). There appeared to be fewer serious adverse events with lopinavir-ritonavir (20%) vs standard of care (32%). In particular, there were fewer cases of acute respiratory distress syndrome with lopinavir-ritonavir compared to standard of care (13% vs 27%). Limited data were available for comparison of lopinavir-ritonavir to other treatments.

**Conclusions:** Based on currently available data, there was no clear benefit for use of lopinavir-ritonavir compared to standard of care in severe COVID-19. Risk data suggested a possible decrease in serious adverse events, including acute respiratory distress syndrome. Overall, the benefit-risk profile for lopinavir-ritonavir in severe COVID-19 cannot be considered positive until further efficacy and effectiveness data become available.

## 1 Introduction

Coronaviruses have circulated in human and animal populations for many years and in humans they are a cause of respiratory tract infections [1]. More recently, Severe Acute Respiratory Syndrome coronavirus 2 (SARS-CoV-2) emerged in Wuhan, China in December 2019 [2, 3]. SARS-CoV-2 causes coronavirus disease (COVID-19) and this outbreak was declared a global pandemic by the World Health Organisation (WHO) in March 2020 [4]. Fever, cough and shortness of breath are the main reported symptoms of COVID-19 [5] but this disease also has a concerning case mortality rate among certain populations, such as older adults and those with underlying health conditions.

In the current COVID-19 pandemic, there is a need to identify effective, safe treatments as rapidly as possible. Lopinavir-ritonavir (LPVr) is a combination protease inhibitor and nucleoside analogue, used for the treatment of human immunodeficiency virus (HIV-1) [6]. The use of LPVr in severe acute respiratory syndrome (SARS) has been examined previously and indicated a favourable clinical response [7]. For this reason, multiple trials in COVID-19 are currently being conducted to determine if LPVr is an effective treatment, including the world-wide RECOVERY trial and the WHO’s SOLIDARITY trial [8-10]. It is essential to examine the benefit-risk profile of all medications, but ongoing monitoring is especially important where treatments may be used with limited evidence in new indications. Lopinavir-ritonavir is already being used as a standard treatment for COVID-19 in some countries, although a systematic benefit-risk assessment on the use of LPVr for COVID-19 treatment, based on currently available evidence, has not yet been conducted.

The Benefit-Risk Action Team (BRAT) uses a structured, descriptive framework to outline the key benefits and risks of a medication within a defined disease context. If sufficient relevant data are available, additional quantitative assessment can be used to further examine the benefit-risk profile [11]. BRAT was also designed to assist communication with regulatory authorities [12]. The framework design allows for transparency in the decision making process and assumptions can be explored further by sensitivity analyses using quantitative methods[13].

The systematic benefit-risk assessment for LPVr was conducted based on publicly available data to May 13^th^ 2020. Due to continuous emerging data on the use of LPVr in COVID-19, the framework can be subsequently used to repeat the assessment as further data arise, allowing for rapid and dynamic evidence-based decision-making as more relevant data become available.

## 2 Objectives

To examine the benefit-risk profile of lopinavir-ritonavir in COVID-19 patients compared to standard of care, placebo or other treatments. A key objective of this study was to provide a platform for a dynamic systematic benefit-risk evaluation; although initially this evaluation is likely to contain limited information, it is required due to the urgent unmet public need. Importantly it allows additional data to be incorporated as it becomes available, and re-evaluation of the benefit-risk profile.

## 3 Methods

### 3.1 Benefit-Risk Framework

#### 3.1.1 Population of interest

Patients with COVID-19 who were treated with LPVr were the population of interest, while patients receiving standard of care, placebo or other treatments were the comparator of interest.

#### 3.1.2 Outcomes of interest

Key benefits and risks, i.e. those considered to be of clinical importance or potentially serious, were included in the value tree which provides a visual representation of these outcomes in the context of severe COVID-19 disease. These benefits and risks displayed in the value tree were ranked according to perceived importance (benefits) and potential seriousness (risks). Risks were categorised according to which system organ class (SOC) they belonged to, and where multiple events were identified within the same SOC, the ranking was based on the most serious event(s) within that SOC, with the most serious event(s) in each SOC presented first.

#### 3.1.3 Data sources and customisation of the framework

A literature search was performed in PubMed and Embase using the following search strategy:

(lopinavir AND ritonavir) AND (covid* OR SARS-CoV-2 OR n?CoV OR coronavirus)

Papers were included if they reported quantitative data on effectiveness and/or safety of LPVr in patients with severe COVID-19. Case reports and case series were excluded. Results were restricted to English language only (abstracts in English language were acceptable where sufficient data provided) and peer-reviewed publications since 2019 to 13^th^ May 2020. Data were extracted for each benefit and risk, for LPVr and the comparator (standard of care, placebo or other treatments), where available. EudraVigilance spontaneous reporting data (up to 8th May 2020) for LPVr where used in COVID-19 were also examined.

### 3.2 Outcome assessment

Key benefits and risks associated with the use of LPVr were identified from available data sources, including the product information, regulatory assessment reports, and published literature. Predicted key benefits (clinical endpoints) were derived from both published literature and in the case of ongoing studies, available clinical trial protocols. A summary benefit-risk table was created to allow visualisation of the magnitude of each benefit and risk. Risk differences and corresponding 95% confidence intervals (CI) were calculated for each outcome where both numerator (number of events) and denominator (number of patients at risk) were available for both the treatment group (LPVr) and comparator group. No appropriate comparator groups were identified in EudraViligance for LPVr. Consequently, spontaneous reports are not included in the benefit-risk table and are presented in the text only.

## 4 Results

Figure 1 displays the value tree of the key benefits and risks related to LPVr treatment in COVID-19.

**Fig 1.**
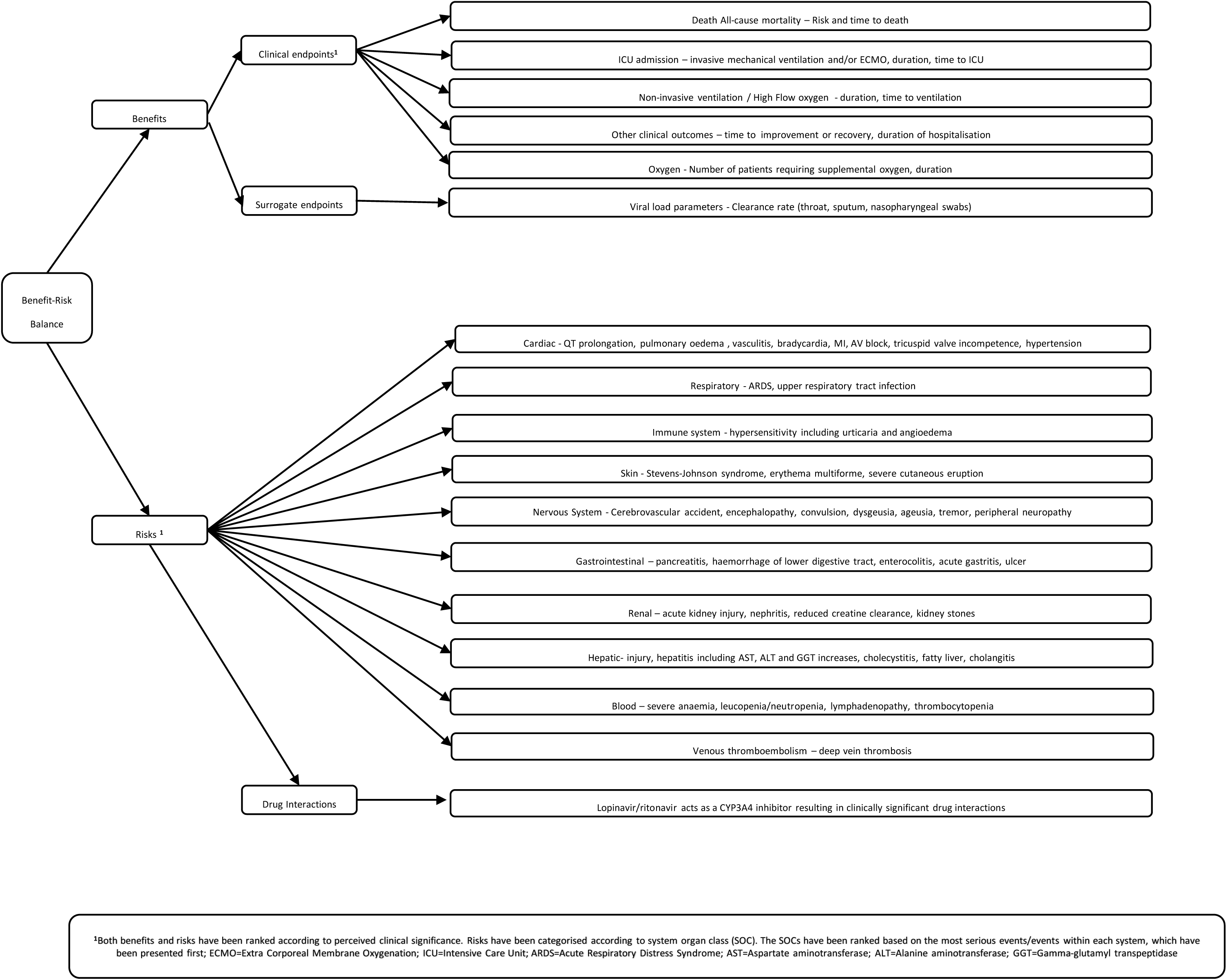
Value tree of key benefits and risks identified for lopinavir/ritonavir, ranked by order of clinical significance.

### 4.1 Benefits

Key benefits have been listed in the value tree in descending order of perceived clinical importance. At the current time, only one clinical trial was identified which provided empirical data for any of the clinical endpoints listed in the value tree [14]. Whilst the primary objective of this clinical trial was time to clinical improvement, additional data was provided for various endpoints including mortality risk, risk and duration of invasive mechanical ventilation, risk of non-invasive ventilation and oxygen requirement.

### 4.2 Risks

Key risks were identified for LPVr based on the current available evidence. It is acknowledged that this product is not licenced for use in the treatment of COVID-19 disease, and whilst safety data is available for its licenced use in HIV-1, its safety profile in the context of COVID-19 is largely unknown. Furthermore, for the limited safety data that is available for its use in COVID-19, it is often unclear whether the reported adverse event is due to the use of LPVr, or attributable to the underlying disease. Potentially serious risks that are likely to still pose a risk with the proposed short-term use of LPVr for COVID-19 have been summarised in the value tree and ranked according to perceived seriousness.

One of the most serious risks is prolongation of the QT interval, and the subsequent increased risk of sudden cardiac death [15-18]. Patients with COVID-19 are already predisposed to the development of cardiac arrhythmia due to the effect of the virus on metabolic dysfunction, myocardial inflammation and the sympathetic nervous system [16]. Also it is important to note that LPVr is an inhibitor of CYP3A4, and therefore it cannot be used with other medicines which are substrates of this enzyme, such as chloroquine, which itself can cause QT prolongation [17]. In addition to the effects on QT prolongation, LPVr has also been shown to cause modest asymptomatic prolongation of the PR interval in some healthy adult subjects, with rare reports of 2^nd^ or 3^rd^ degree atrioventricular block in patients with underlying structural heart disease and pre-existing conduction system abnormalities or in patients receiving drugs known to prolong the PR interval (such as verapamil or atazanavir), and therefore LPVr should be used with caution [19] in such patients.

Lopinavir/ritonavir are both inhibitors of the P450 isoform CYP3A, and therefore treatment is likely to increase plasma concentrations of concomitant medicinal products that are primarily metabolised by CYP3A [19]. Clinically significant drug interactions have been observed with LPVr use during treatment for COVID-19, including increased plasma levels of direct oral anticoagulants [20], and increased plasma levels of immunosuppressants in organ transplant recipients [21].

Certain risks factors and clinical characteristics have been associated with the development of acute respiratory distress syndrome (ARDS) amongst patients with COVID-19 [22]. Patients who developed COVID-19 related ARDS were likely to require admission to the intensive care unit (ICU). In the study by Cao et al, respiratory failure/ARDS was reported as a serious adverse event in both treatment groups [14]. Whilst causality in these cases is not known, it would seem likely that these cases were attributable to progression of the underlying COVID-19 disease.

Hypersensitivity reactions such as urticaria and angioedema are reported to occur commonly with the use of LPVr for the treatment of HIV, and rarely serious skin reactions such as Stevens-Johnson syndrome and erythema multiforme have been reported with its use in this treatment population [19]. Gastrointestinal side effects of LPVr are well recognised, and diarrhoea and nausea are very common. Serious gastrointestinal adverse effects included in the key risks for this assessment include pancreatitis, which has been associated with the use of LPVr [19, 23, 24]; most patients who developed pancreatitis during treatment for HIV had a prior history of this condition. Treatment with LPVr has been associated with an increase in triglycerides in patients treated for HIV [19], and amongst patients treated for COVID-19 [25-28], which is likely to be another contributory factor in the development of pancreatitis.

In the context of treatment for HIV, LPVr has been uncommonly associated with certain adverse renal outcomes, including a reduction in creatine clearance, nephritis and haematuria [19]. Cases of acute kidney injury have been reported in patients taking LPVr in COVID-19, however it is unclear whether there is any association, as this outcome was reported more frequently amongst patients in the standard of care comparator group [14] in addition to overall limited safety data availability. Elevations of liver enzymes have also been commonly reported with the use of LPVr in the treatment of HIV [19], and liver injury has been reported in patients treated with LPVr for COVID-19 [29, 30]. Blood dyscrasias have also been associated with the use of LPVr during HIV treatment [19], with reports of severe anaemia amongst patients treated for COVID-19 [14].

### 4.3 Quantitative data

Data for outcomes are presented in the data extraction table and key benefit-risk summary table (Tables 1 and 2, respectively). From literature searching we identified 143 papers from PubMed and 264 papers from Embase for LPVr. After initial review and removal of duplicates, 15 papers were reviewed further to determine whether they met all inclusion criteria; seven papers were included in the final benefit-risk assessment.

**Table 1.**
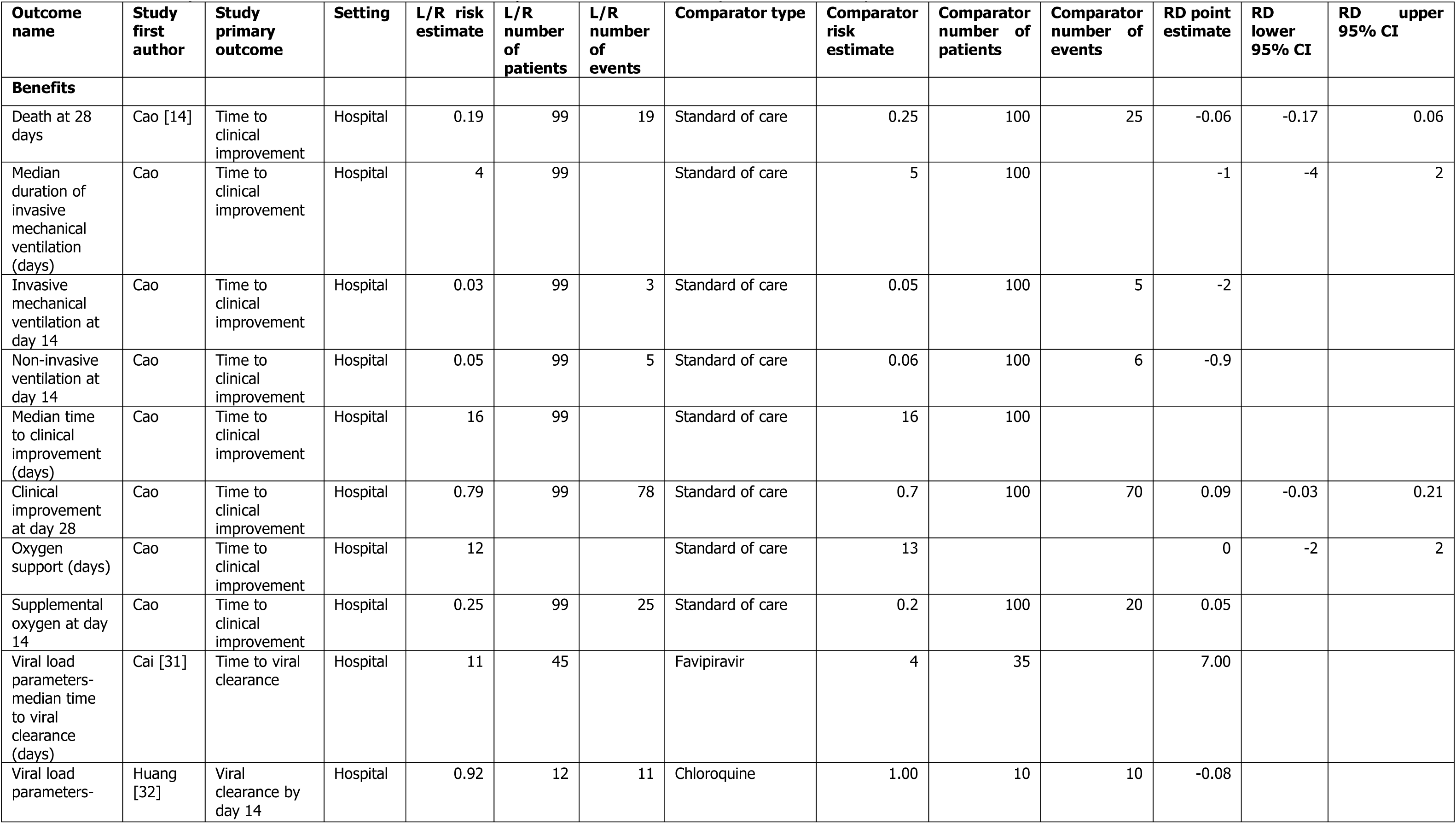
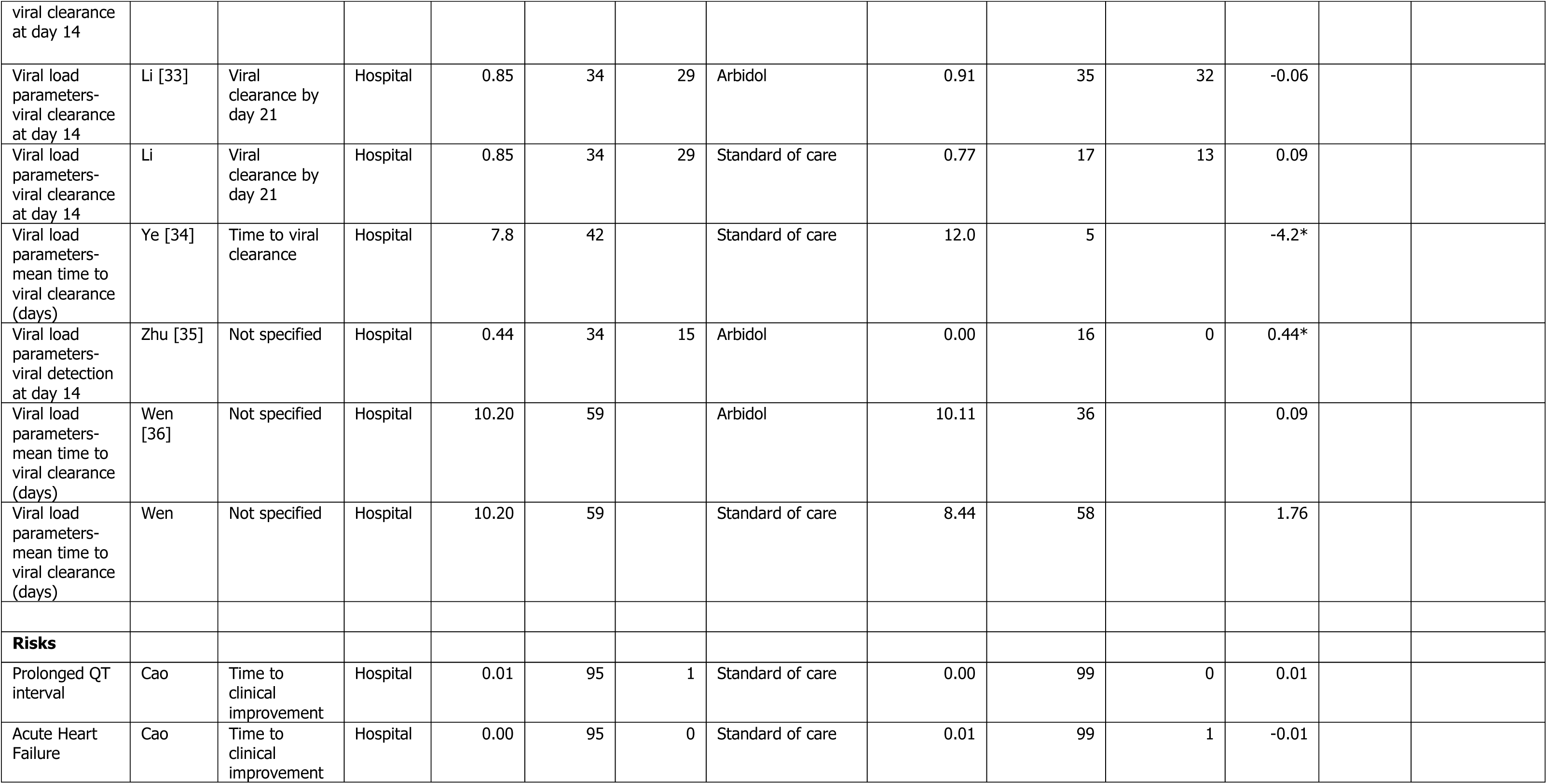
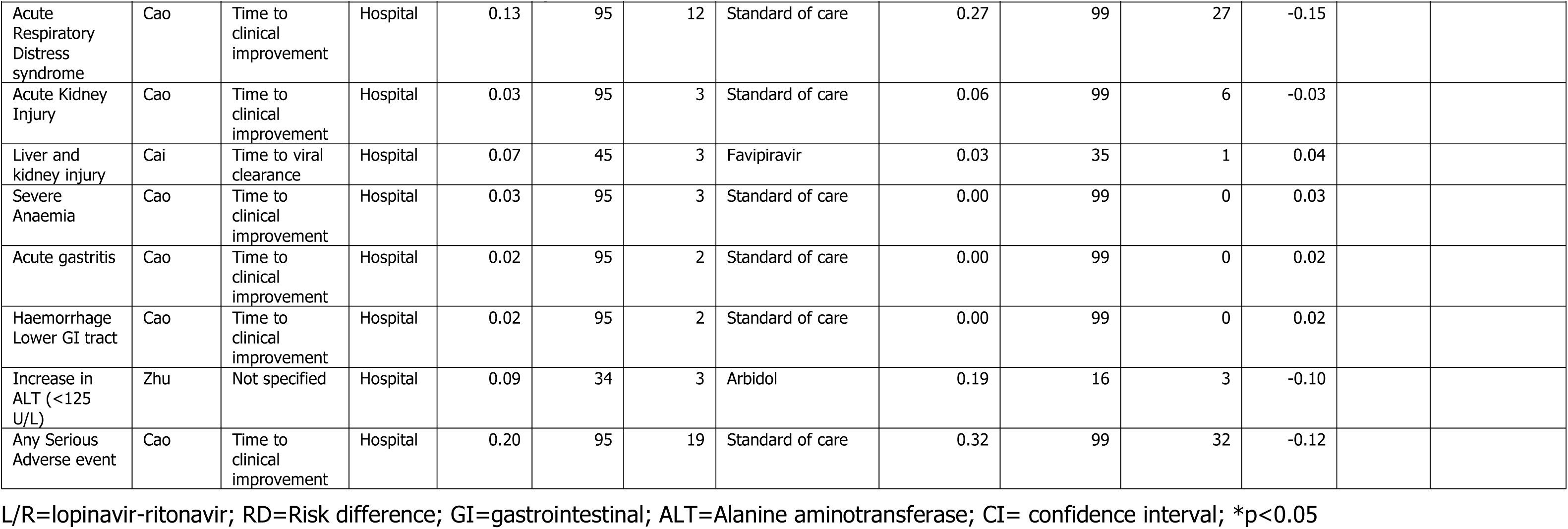
Data for key benefits and risks identified for lopinavir-ritonavir from peer-reviewed, published literature.

**Table 2.** Benefit-Risk summary table for key benefits and risks identified for lopinavir-ritonavir compared to standard of care.

| Outcome name | L/R risk/1000 patients | Standard of care risk/1000 patients | RD (95% CI)/1000 patients | Hazard ratio (95% CI) |
| --- | --- | --- | --- | --- |
| **Benefits** |  |  |  |  |
| Death at 28 days | 192 | 250 | -58 (-173, 57) |  |
| Invasive mechanical ventilation at day 14 | 30 | 50 | -20 |  |
| Non-invasive ventilation at day 14 | 51 | 60 | -9 |  |
| Time to clinical improvement |  |  |  | 1.31 (0.95, 1.80) |
| Clinical improvement at day 28 | 788 | 700 | 88 |  |
| Supplemental oxygen at day 14 | 253 | 200 | 53 |  |
| Viral load parameters- viral clearance at day 21 | 853 | 765 | 88 |  |
| **Risks** |  |  |  |  |
| Prolonged QT interval | 11 | 0 | 11 |  |
| Acute Heart Failure | 0 | 10 | -10 |  |
| Acute Respiratory Distress syndrome | 126 | 273 | -147 |  |
| Acute Kidney Injury | 32 | 61 | -29 |  |
| Severe Anaemia | 30 | 0 | 30 |  |
| Acute gastritis | 21 | 0 | 21 |  |
| Haemorrhage Lower GI tract | 20 | 0 | 20 |  |
| Any Serious Adverse event | 200 | 323 | -123 |  |

L/R=lopinavir-ritonavir; RD=Risk difference; CI= confidence interval; GI= gastrointestinal

In comparison to standard of care, data for several key benefits and risks for LPVr were identified. In the Cao et al trial [14], the benefit of time to clinical improvement (intention to treat analysis) was not statistically significant after adjustment for other covariates in comparison to standard of care (16 vs 16 days, HR=1.31, 95% CI: 0.95, 1.80). Other non-significant benefit data were identified in this trial including median duration of mechanical ventilation (RD=-1 days, 95% CI: -4, 2) and death at 28 days (RD=-0.06, 95% CI: -0.17, 0.06).

Risk data were mainly available from the Cao et al trial, which reported fewer serious adverse events in patients taking LPVr (20%) compared to standard of care (32%). There were fewer cases of ARDS with LPVr compared to standard of care (13% vs 27%).

Limited data were available for comparison of LPVr to other treatments. Viral clearance at day 14 for LPVr was lower compared to arbidol (85% vs 91%), and there was minimal difference in mean time to viral clearance (10.20 days vs 10.11 days, respectively). Data were only available for one risk in comparison to arbidol; a lower proportion of those patients treated with LPVr experienced increased ALT compared to arbidol (9% vs 19%, respectively).

In spontaneous reports from Eudravigilance, for LPVr used in COVID-19, there were 28 reports of hepatocellular injury, 15 reports of acute kidney injury, 17 reports of prolongation of the QT interval, six reports of ARDS and three reports of pancreatitis. Information on case reports was only available from the publicly available dataset and so was limited.

## 5 Discussion

This paper provides a systematic benefit-risk assessment using the BRAT methodology and is inclusive of available literature up to and including 13^th^ May 2020. Therefore, this represents a snapshot of the data available to date and outlines a clear and transparent framework into which subsequent clinical trial and observational study data can be incorporated, and the benefit-risk profile re-assessed.

At the current time, data relating to the benefits of LPVr are limited, with efficacy data available from only one published clinical trial. This trial found no statistically significant difference in the primary outcome of time to improvement (intention to treat analysis) between the two groups (LPVr treatment in addition to standard supportive care vs. standard care alone), however the sample size was small. There was some evidence (though not statistically significant) that LPVr reduced mortality at 28 days [95% CI: 19.2% vs. 25.0%, difference of -5.8% (95% CI: -17.3% to 5.70%)]. It is of note that the median time interval between symptom onset and randomization was 13 days (IQR, 11 to 16 days), therefore it is unknown whether more favourable results may have been seen if drug treatment had been initiated earlier in the course of the disease.

The safety profile of LPVr in the treatment of severe COVID-19 disease is largely unknown. We identified key risks from its usage in the treatment of HIV, in addition to the limited safety data available from its use in COVID-19. Comparator safety data revealed a lower incidence of both ARDS and all serious adverse events amongst patients receiving LPVr compared to standard of care ([risk difference -147 events per 1000 patients], [-123 events per 1000 patients], respectively). Very small numbers of cases of other adverse events were reported in the study by Cao et al., including severe anaemia, acute gastritis and lower gastrointestinal bleeding, all of which were only reported amongst patients taking LPVr. The incidence of acute kidney injury was lower in the Cao et al trial amongst patients taking LPVr compared to standard of care (3% vs 6%).

Overall, there was a lack of efficacy data with no clear benefits identified for LPVr treatment compared to standard of care. Risk data, although limited, suggested a possible decrease in adverse events for some serious outcomes compared to standard of care. Further data is needed on efficacy and effectiveness of LPVr for severe COVID-19.

### 5.1 Strengths and Limitations

A strength of this approach is the inclusion of all key benefits and risks in the same model and a transparent framework into which further data can be included as and when this becomes available. The method has a significant advantage compared to systematic reviews which are equally comprehensive but focus only on efficacy. When sufficient data is available, the methodology allows benefits and risks to be ranked, and weightings applied based on this ranking, with further quantitative analysis. The reproducibility of this assessment allows multiple treatments to be assessed using this approach, thereby allowing direct comparison between different treatments. This is of great significance during the current COVID-19 crisis, in which several potential interventions currently under evaluation need to be assessed and evaluated in real time, and where new data needs to be incorporated quickly. Regulatory decision makers are also familiar with this framework, facilitating interpretation.

A limitation of the benefit-risk assessment presented at this time is the relative paucity of data that has been included in the model, as many clinical trials assessing LPVr are still ongoing. In addition, trials for which data were available had very small sample sizes and were likely to be underpowered when examining non-primary outcomes. Study quality was also not considered in the assessment, although we only included peer-reviewed manuscripts.

### 5.2 Conclusions

Based on currently available data, there was no clear benefit for use of LPVr compared to standard of care in severe COVID-19. Risk data suggested a possible decrease in serious adverse events, including ARDS. Overall, the benefit-risk profile for LPVr in severe COVID-19 cannot be considered positive until further efficacy and effectiveness data become available.

## Data Availability

Data used in this analysis are available from the references supplied.

## Declarations

### Ethics approval

This study was conducted in accordance with international ethical guidelines. Ethics approval was not required for this study.

### Availability of data and material (data transparency)

Data used in this analysis are available from the references supplied.

### Contributorship statement

VO assisted with study design and data extraction, wrote the study proposal and the first draft of the manuscript. MD assisted with study design, identified outcomes of interest, constructed the value tree and assisted with data extraction. SD assisted with value tree and rankings. DR, SL, JD and AE assisted with study design, literature searching and data extraction. SAWS assisted with the concept, study design and manuscript revisions. All authors reviewed, contributed to revisions and approved the manuscript and accept full responsibility for its overall content.

